# Modes of transmission of COVID-19 outbreak- a mathematical study

**DOI:** 10.1101/2020.05.16.20104315

**Authors:** Chandan Maji

## Abstract

The world has now paid a lot of attention to the outbreak of novel coronavirus (COVID-19). This virus mainly transmitted between humans through directly respiratory droplets and close contacts. However, there is currently some evidence where it has been claimed that it may be indirectly transmitted. In this work, we study the mode of transmission of COVID-19 epidemic system based on the susceptible-infected-recovered (SIR) model. We have calculated the basic reproduction number *R*_0_ by next-generation matrix method. We observed that if *R*_0_ *<* 1, then disease-free equilibrium point is locally as well as globally asymptotically stable but when *R*_0_ > 1, the endemic equilibrium point exists and is globally stable. Finally, some numerical simulation is presented to validate our results.

## 1. Introduction

Today an invisible emperor of the world has put human civilization in front of a big question mark which is popularly known as COVID-19 (World Health Organization (WHO)). Mankind, proud of being the most advanced intelligent creature in the world is now endangered by its tyranny. COVID-19 is an abbreviated form of Severe Acute Respiratory Syndrome Coronavirus 2 (SARS-CoV-2), which has not been previously identified in humans [1]. On Dec, 2019, this disease was first identified in Wuhan city Hubei province of china [2], after that rapidly it has spread out all over the world. SARS [3] and MERS [4], these two previously known corona virus disease outbreaks have already occurred in the last two decades. Till now it is reported that COVID-19 has been spread out in more than 200 countries and now it has taken the shape of an epidemic in worldwide with more than 3.8 million people infected and 2.5 lakhs death [5].

It is presumed that COVID-19 is directly transmitted through human to human via respiratory droplets among closed contact and infected people generally develop signs and symptoms, including mild respiratory discomfort and fever, on an average of 5-6 days after infection [6]. In [7], Jiang et al. estimated that fatality rate of this virus is near about 4.5% but for the age group 70-79 it has gone up to 8%. This disease is more critical for elder people who has other diseases like diabetes, asthma, cardiovascular disease [6]. However, these transmission modes do not explain all cases. A lot of evidences have shown that COVID-19 has highly similar biological properties with severe acute respiratory syndrome coronavirus (SARS-CoV). In their study [8], Cai et al. investigated a cluster of COVID-19 cases associated with a shopping mall in Wenzhou, China and showed that indirect transmission of the causative virus occurred from virus contamination of common objects, virus aerosolization in a confined space, or spread from asymptomatic infected persons. Zhang et al. [9], suggested that transmission may also occur through fomites in the immediate environment around the infected person. Guo et al. [10], tested surface and air samples from an intensive care unit (ICU) and a general COVID-19 ward at Huoshenshan Hospital in Wuhan, China and found that contamination was greater in ICU than general wards, they also have seen that virus was widely distributed on floors, computer mice, trash cans, and sickbed handrails. Therefore, it can be concluded that the transmission of the COVID-19 virus can occur by direct contact with infected people and indirect contact with surfaces in the immediate environment or with objects used on the infected person. Considering direct transmission mode, already a lot of works have been done mathematically and numerically to give an efficient prediction on COVID 19 outbreaks [11, 12, 13, 14, 15]. Researchers are trying their best to discovering vaccines and treatments for the virus but it may be far away from our imagination [16]. In this work, we have formulated a mathematical model of direct and indirect transmission to study the dynamics of COVID-19 outbreaks.

The paper is organized as follows: In Section 2. we have formulated a mathematical model for the epidemic COVID-19. Boundedness of the solutions of system (1) is performed in Section 3. Stability analysis of disease-free equilibrium point is discussed in Section 4. Local and global stability of endemic equilibrium point is presented in section 5. Sensitivity analysis and some numerical simulations are discussed in Section 6, to validate our results. A brief discussion is given in Section 7.

## 2. Model Formulation

The world has now paid a lot of attention on the outbreak of novel coronavirus (COVID-19) which has been declared a pandemic by WHO. Till date, no vaccine or medicine is available to cure the disease properly so people are getting in a panic and they are afraid of disease transmission. According to current evidence, COVID-19 virus is primarily transmitted between people through respiratory droplets and close contacts [17, 18, 19, 20, 21, 22]. But there are also some evidences who conclude that transmission may also occur through fomites in the immediate environment around the infected person [9, 23, 24]. Inspired by above all these work, we have formulated a epidemic model for COVID-19 of direct and indirect transmission. The model is based on traditional susceptible-infected-recovered (SIR) models of disease transmission in humans. Our proposed model is described by a system of four ordinary differential equation which is as follows:

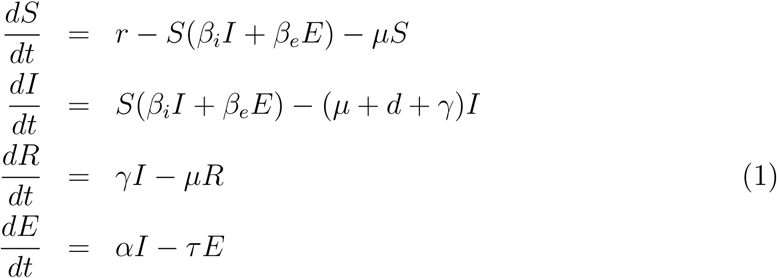

with initial conditions *S*(0) > 0, *I*(0) > 0, *E*(0) ≥ 0, *R*(0) ≥ 0.

Here, *S*(*t*), *I*(*t*) denotes the density of susceptible population, infected population. *R*(*t*) is the total number of recovered population and *E*(*t*) is the mass of infectious material present in the environment. All other model parameters and their description are given in Table 1.

**Table 1.** Parameters and their descriptions

| Table 1. Parameters and their descriptions |  |
| --- | --- |
| $r$ | recruitment rate of susceptible which includes immigrants and newborns |
| $\beta_i$ | direct disease transmission rate |
| $\beta_e$ | indirect disease transmission rate |
| $\mu$ | natural death rate |
| $d$ | disease induced death rate |
| $\gamma$ | recovery rate from infected population |
| $\alpha$ | infectious material in the environment |
| $\tau$ | loss of infectious material from the environment |

## 3. Boundedness

For biological validity of system (1), it is necessary to prove that all solutions of system (1) with positive initial values will remain positive for all time *t >* 0. Thus in this section we want to prove the positivity and boundedness of solutions of our considered system.

### Lemma 1

All solutions (*S*(*t*),*I*(*t*),*R*(*t*),*E*(*t*)) of system (1) with positive initial values in 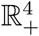 will remain positive for all *t* > 0

#### Proof

From first equation of system (1), we get

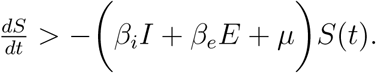

Thus, 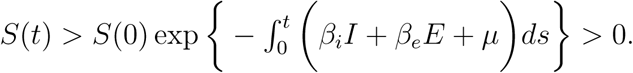.

As, *S*(0) > 0 then *S*(*t*) > 0 for all *t* > 0.

Similarly, it can be shown that *I*(*t*) > 0, *R*(*t*) > 0, *E*(*t*) > 0 for all *t* > 0. Hence the interior of 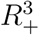 is an invariant set of system (1).

### Lemma 2

All solution of system (1), which initiate in 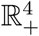 are bounded and lie in the region 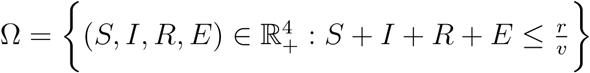.

#### Proof

Define a function, 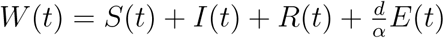.

The time derivative along the solution of system (1), we get

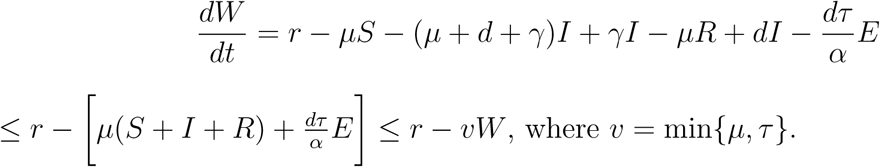

By using differential inequality argument [25], we get

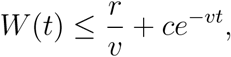

where *c* is arbitrary positive constant. Hence we get, 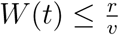 when *t* → ∞.

All solution of system (1) enter into the region 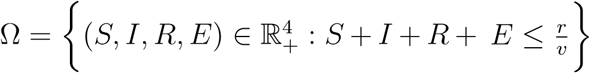.

## 4. Stability analysis

Basic reproduction number is one of the most important threshold parameter which can determine whether the infectious disease will die out or spread through population with time increases. Here we calculate the basic reproduction number for COVID-19 model through next generation matrix method [26].

System (1) has a unique disease free equilibrium point *E*_1_(*K*, 0, 0, 0), where 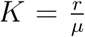. Then using the notation in [26], the reproduction number *R*_0_ of system (1) is given by

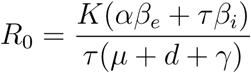

### Theorem 1

Disease free equilibrium *E*_1_ is locally as well as globally asymptotically stable if *R*_0_ < 1 and unstable if *R*_0_ > 1.

#### Proof

The Jacobian matrix at *E*_1_ is given by

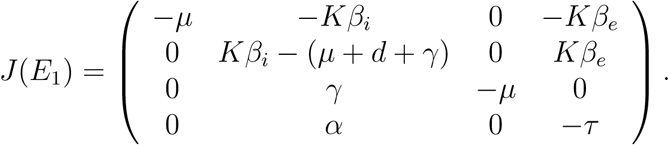

Therefore, all eigenvalues of the characteristic equation of *J*(*E*_1_) are −*μ*, −*μ* and other two eigenvalues are the roots of the equation

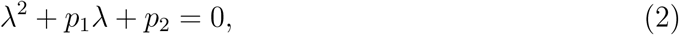

where, *p*_1_ = (*μ* + *d* + γ + *τ* − *Kβ_i_*), *p*_2_ = *τ*(*μ* + *d* + γ) − *K*(*αβ_e_* + *τβ_i_*).

Clearly, *p*_1_,*p*_2_ > 0 when *R*_0_ < 1. Therefore all roots of equation (2) has negative real part, hence *E*_1_ is locally asymptotically stable and unstable when *R*_0_ > 1.

Now, we consider the following Lyapunov function to prove global stability of disease free equilibrium point *E*_1_.

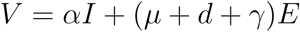

with Lyapunov derivative we obtain,

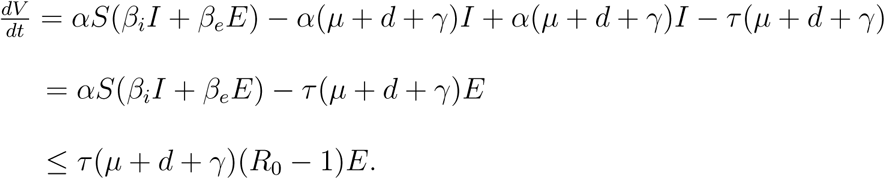

Since, all parameters of the model are non negative. Hence, it follows that 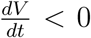 when *R*_0_ < 1, hence by Lasalle invariance principle [27], the disease free equilibrium point is globally asymptotically stable.

## 5. Endemic equilibrium and its stability analysis

The endemic equilibrium point 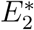 of the model (1) is given by

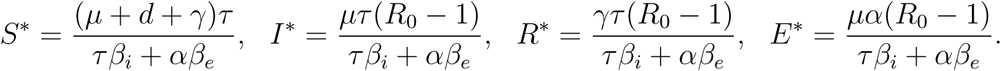

Clearly, we have seen that the endemic equilibrium point *E*^*^ is feasible if *R*_0_ > 1.

### Theorem 2

Endemic equilibrium point 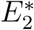 is locally asymptotically stable if *R*_0_ > 1

#### Proof

The Jacobian matrix of system (1), at endemic equilibrium point 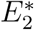 is given by

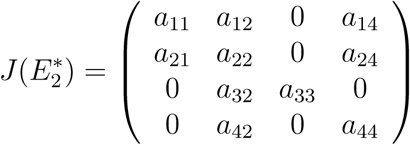

where a_11_ = −*μR*_0_, 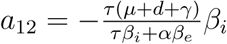, *a*_13_ = 0, 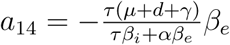, *a*_21_ = *μ* (*R*_0_ − 1), 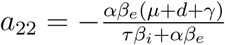, *a*_23_ = 0, 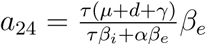 *a*_31_ = *a*_34_ = *a*_41_ = *a*_43_ = 0, *a*_32_ = γ, *a*_33_ = −*μ*, *a*_31_ = *α*, *a*_44_ = −*τ*..

Characteristic equation of 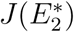 is given by

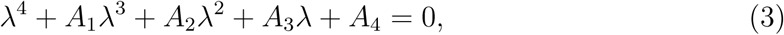

where, *A*_1_ = − (*a*_11_ + *a*_22_ + *a*_33_ + *a*_44_),

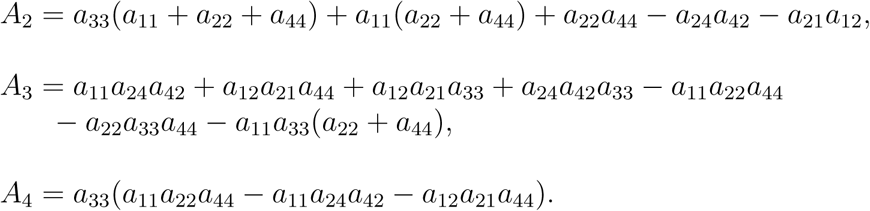

It follows from Routh-Hurwitz criteria [28] that all roots of equation (3) will have negative real parts if *A*_1_ > 0, *A*_3_ > 0, *A*_4_ > 0 and *A*_1_*A*_2_ > *A*_3_, 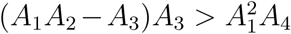. This will be possible when *R*_0_ > 1. Hence, endemic equilibrium point 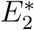 is locally stable.

System (1) is said to be uniformly persistent if there exist a constant *ā* such that any solution of (1) satisfies

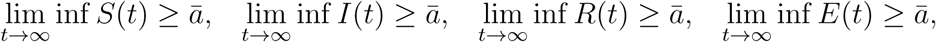

From Theorem 1, we get the disease free equilibrium *E*_1_ is unstable for *R*_0_ > 1. Now apply the uniform persistence result defined in [29], and then using the approach used to prove Proposition 3.3 of [30], it can be proved that system (2) is uniformly persistent in the region Ω.

### Theorem 3

Assume, *R*_0_ > 1, then endemic equilibrium point 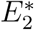 is globally asymptotically stable if the following conditions hold:

(i) 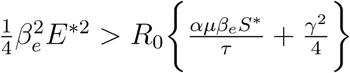 and det(*M*) > 0.

#### Proof

Consider a positive definite function

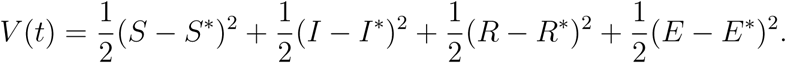

Now differentiating *V* with respect to *t* along the solutions of system (2), we get

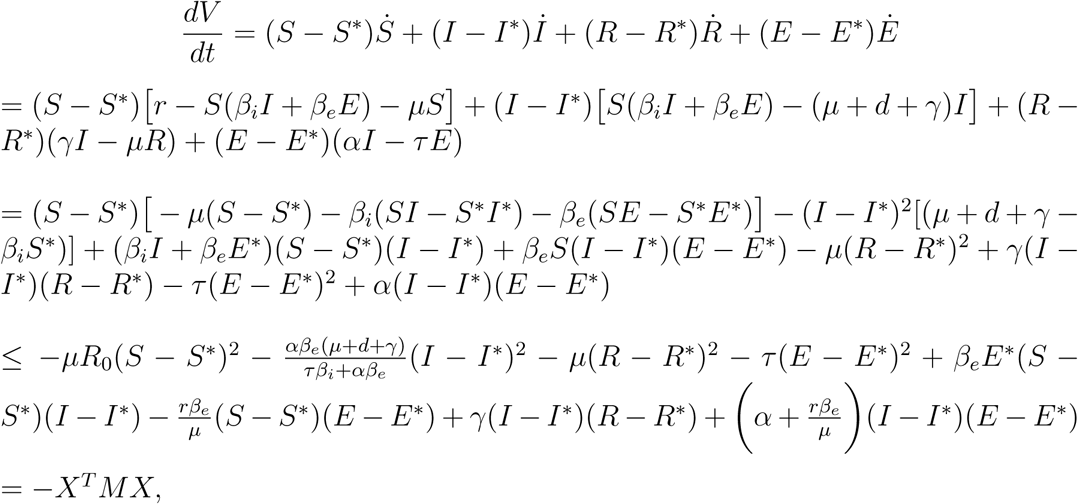

where *X^T^* = {|*S* − *S*^*^|, |*I* − *I*^*^|, |*R* − *R*^*^|, |*E* − *E*^*^|} and *M* = (*a_ij_*)_4×4_. Elements of the matrix *M* are given by

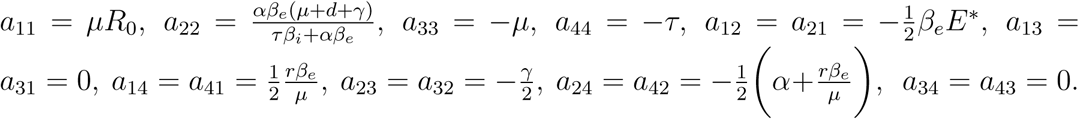

Therefore, *M* is positive negative if (i) 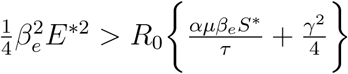 and det(*M*) > 0.

Thus, 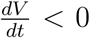 and consequently *V* is a Lyapunov function and hence endemic equilibrium point *E*^*^ is globally asymptotically stable.

## 6. Sensitivity Analysis and Numerical simulation

Sensitivity analysis is one of the most important part to get an overview of most influential parameters in modelling of a infectious disease. As system stability determined by its reproduction number (*R*_0_), so we want to verify how the sensitive parameters are related with *R*0 and therefore we compute the sensitivity analysis of *R*_0_ with respect to the model parameters. By the definition of the normalized sensitivity index of *R*_0_ with respect to *β* is given by

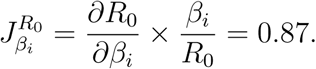

The sensitivity indices of *R*_0_ with respect to other parameters are as follows:

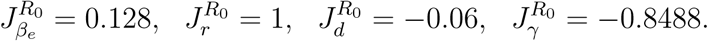

From this analysis we observed that, 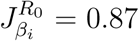, 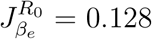, which implies if the direct and indirect disease transmission rate increase by 1% then it will increase the value of *R*_0_ by 0.87% and 0.128% respectively. Again, we see that 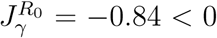 that means increase the value of γ by 1% then the value of *R*_0_ will be reduced by 0.84%. Thus, it can be concluded that reproduction number is positively corelated with *β_i_* and *β_e_* and negatively corelated with γ.

**Figure 1:**
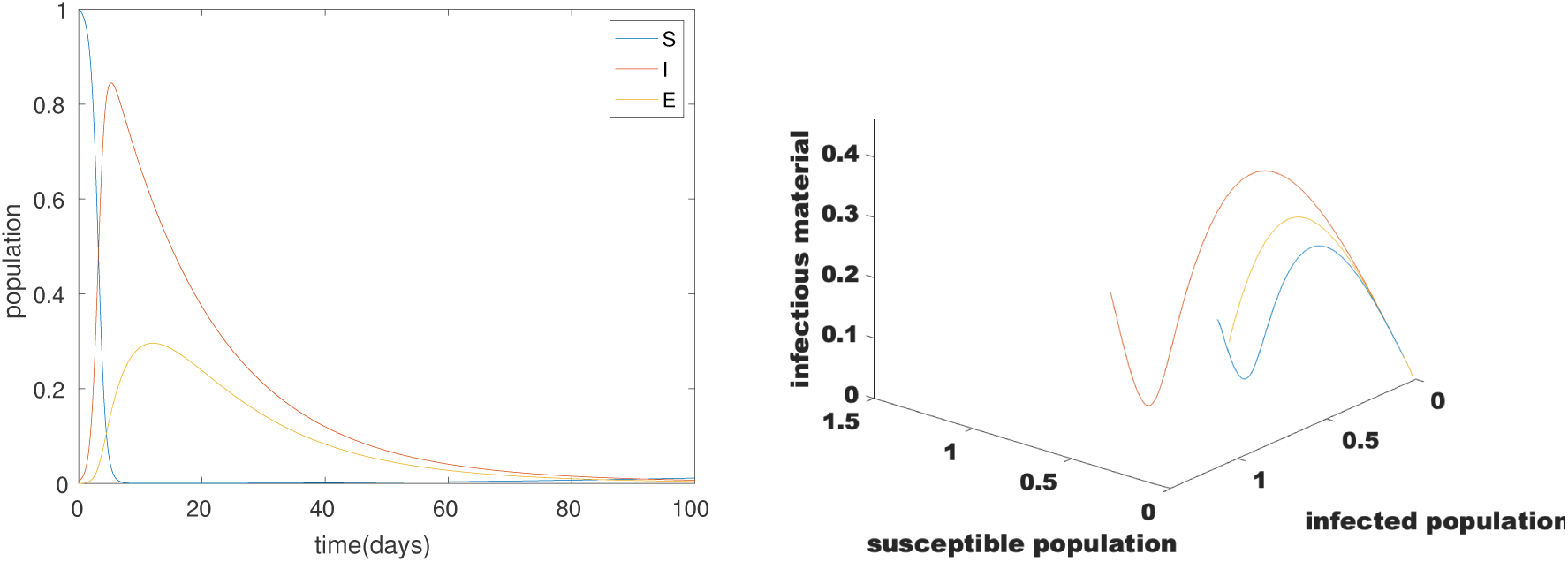
Dynamics of all population over time (days), and endemic equilibrium point is globally stable

To observe the dynamics of our proposed model through numerically we set the following set of parameters hypothetically: *r* = 0.00045308; *β_i_* = 1.7 [5]; *β_e_* = 0.5 [5]; *α* = 0.1; *μ* = 0.005; γ = 0.05; *τ* = 0.2; *d* = 0.0039 [22]. For this set of parameters, we obtain *R*_0_ = 3.1794 > 1, which satisfy the conditions of Theorem 3 and consequently the endemic equilibrium point is globally stable (Figure 1). From this figure we have seen that, susceptible population is decreasing but infected population is increasing over time that means more people are being infected within a short time period and as a result there is an outbreak within a short time span.

Till date, no vaccine or medicine is available to cure the disease properly so to prevent the disease transmission it is necessary to obey social distancing or lockdown which have already been applied by all Countries. The effectiveness of lockdown of 21 days or more days is described in Figure 2. From this figure, we observed that 21 days’ lockdown is not sufficient to control the disease but if we would applied 54 days’ lockdown or more then it shows that the lowest number of people would be infected and the epidemic might be under control. In Figure 3, we observed that how infected population is related with disease transmission rate.

**Figure 2:**
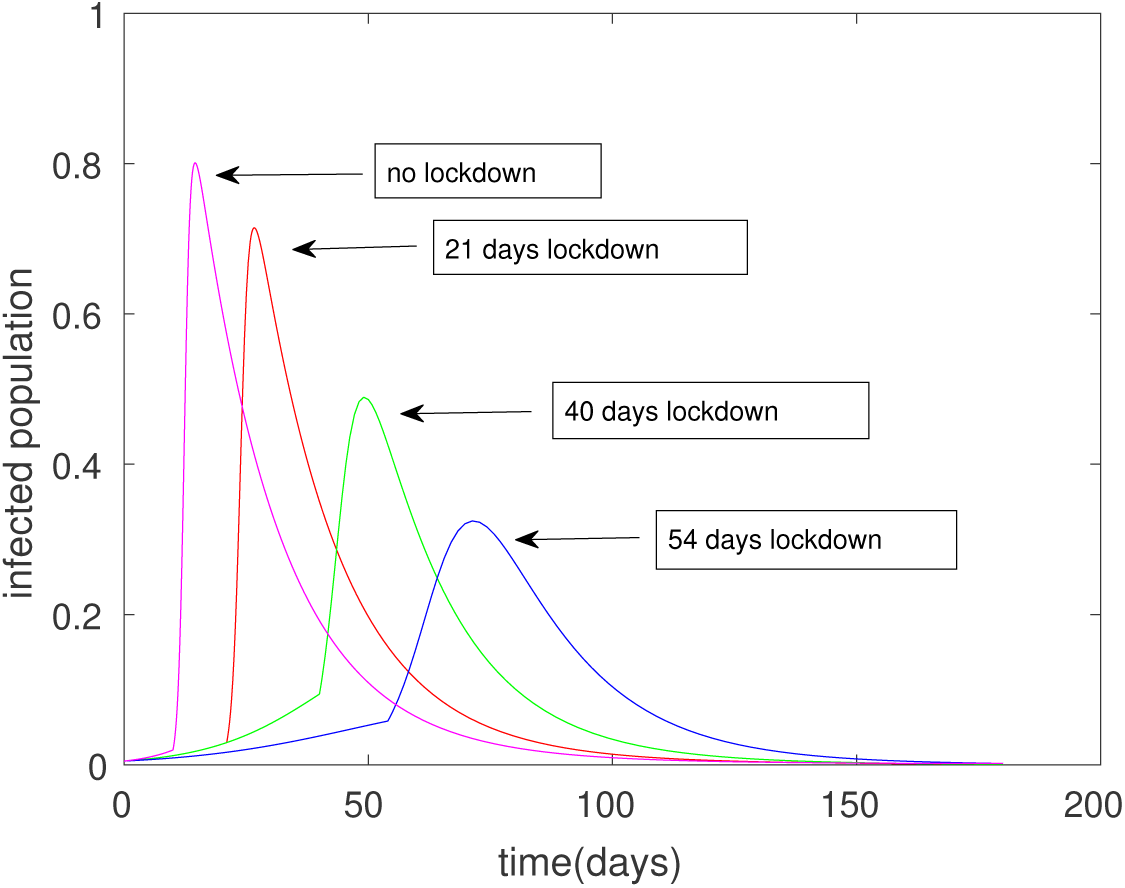
Infected population for different disease transmission rate

**Figure 3:**
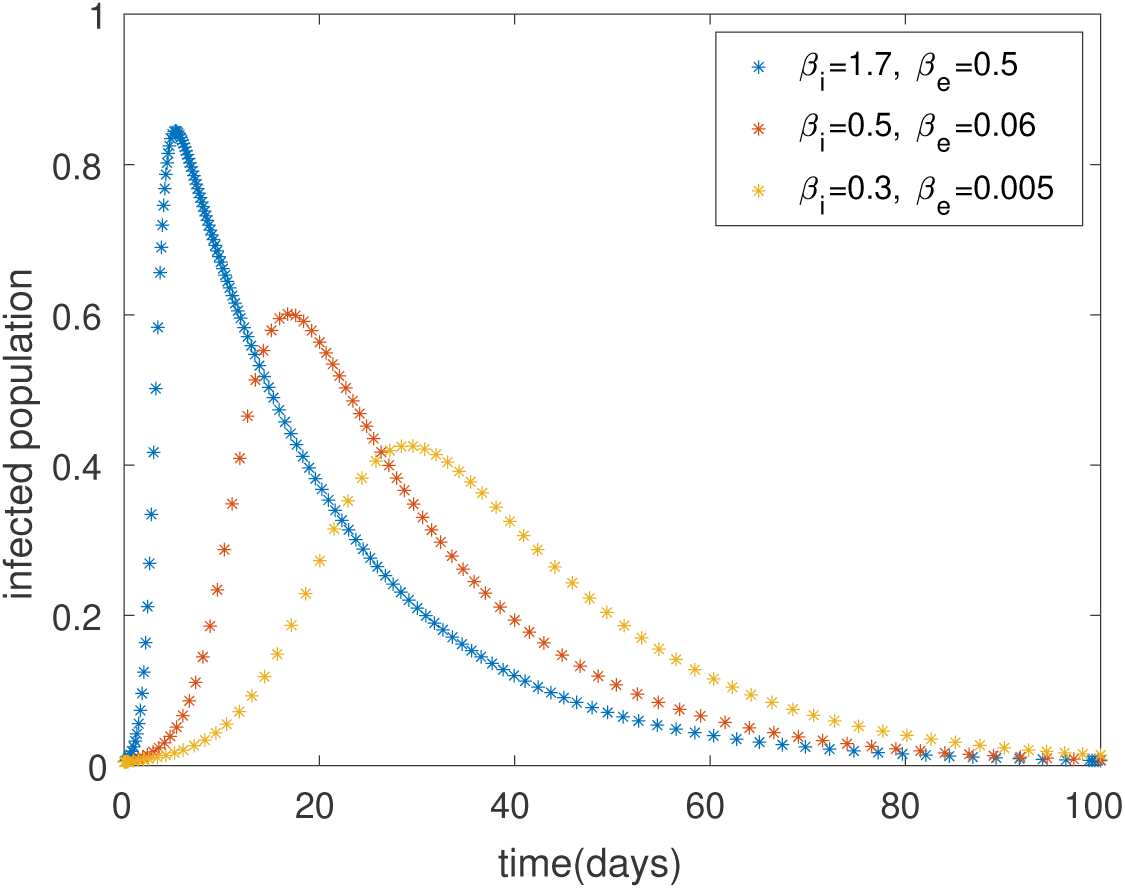
Dynamics of all population over time (days), and endemic equilibrium point is globally stable

## 7. Discussion

Mathematical modelling is one of the best way to express the dynamics of an epidemic. In this paper, we have developed a mathematical model considering the direct and indirect disease transmission rates for COVID-19 outbreaks. The basic reproduction number for our considered model is given by

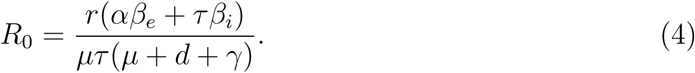

The asymptotic behaviour of our model is determined by it’s basic reproduction number. We observed that if *R*_0_ < 1, then the disease-free equilibrium is globally asymptotically stable. If *R*_0_ > 1, then disease persists in the system and endemic equilibrium point *E*^*^ is globally asymptotically stable (see Figure 1.). From equation (9), we have also seen that the basic reproduction number is proportional to the recruitment rate of the susceptible population and directly as well as indirectly disease transmission rate, that means the probability of disease transmission is higher among the close contact or indirect contact with surfaces in the immediate environment or with objects used on the infected person. From sensitivity analysis it is noticed that basic reproduction number is positive corelated with *β_i_* while negatively corelated with *γ*. Thus to control the disease we must keep the basic reproduction number below the unity.

## Data Availability

There are some hypothetical data and other data has been referred.

## Conflict of interest

The author declare that there is no conflict of interest.

## Notes

### Competing Interest Statement

The authors have declared no competing interest.

### Funding Statement

No funding available.

